# The Multiple Layers of Childhood Adversity and Mortality: Population-based Study Synthesizing Life-course Data on Individual, Family and Neighborhood Adversity in 1.2 million individuals

**DOI:** 10.1101/2025.10.27.25338857

**Authors:** NH Rod, SK Bennetsen, LK Elsenburg, CE Sabel, D Taylor-Robinson, D Kovacs, AG Zucco, TR Vries

## Abstract

**Background:** Childhood adversity is multi-layered, extending beyond the family to include broader neighborhood and health contexts.

**Methods:** Children were followed from birth into young adulthood (16-42 years) using nationwide register data on multi-layered childhood adversity and mortality. Individual adversity included perinatal adversity (preterm or small-for-gestational-age) and mental and physical health service use. Family adversity included five distinct groups using group-based multi-trajectory modelling based on 12 adversities. Neighborhood adversity included material deprivation in small-area geographical zones. We evaluated associations of these layers with all-cause mortality using survival analyses.

**Findings:** 1,235,519 individuals born between 1980 and 2001, were followed up until Dec 31, 2022, capturing 7,320 deaths. Children facing high family adversity were more likely to have perinatal adversity, use health services, and live in deprived neighborhoods. Each layer separately predicted mortality, with for example high physical health service use (HR: 2.36; 95% CI: 2.24; 2.48) and living in a deprived neighborhood (HR: 1.20; 95% CI: 1.14; 1.26) being associated with higher mortality. Cross-layer interactions were most pronounced between family adversity and child health or perinatal adversity. The highest mortality risk was observed among those with both high family and individual adversity: HR: 7.16 (95% CI: 6.40; 8.01) compared to those with low adversity.

**Interpretation:** The co-occurrence and interaction of childhood adversities across different layers can create highly vulnerable groups, deepening lifelong health inequalities. This underscores the importance of a comprehensive, multi-layered approach that targets individual vulnerabilities as well as the broader social environment.

**Funding:** The European Research Council.

**RESEARCH IN CONTEXT:** *Evidence before this study:* We searched for relevant studies in PubMed with no language restrictions from inception to March 31, 2025, using the terms ((“Childhood adversity” OR “Adverse Childhood Experiences” OR “Early life adversity OR “Perinatal” OR Neighbo*rhood deprivation”) and (“[mortality] OR “premature mortality” OR “death” OR “suicide”)”. We identified studies investigating how individual forms of childhood adversity contribute to the risk of mortality in young adulthood, but only a limited number of studies investigating intersections of different forms of adversity in smaller samples. We also identified several conceptual frameworks that have emphasized the importance of considering multiple, intersecting forms of adversity across developmental contexts. Empirical studies that comprehensively investigate the intersections of multiple forms of adversity simultaneously remain limited.

*Added value of this study:* By leveraging large-scale, multi-layer life-course data, the study examines emergent patterns of childhood adversity arising from co-occurring and interacting adversities across individual, family, and neighborhood layers, and relate this to mortality in young adulthood. We demonstrate that adversity is not confined to isolated domains: children exposed to family adversity are more likely to experience perinatal complications, health difficulties, and residence in deprived neighborhoods. Importantly, we show that these adversities are not only independently associated with mortality but can also interact across layers, where adversity at one level amplifies the harmful effects of adversity at another. Such cross-layer interactions help identify highly vulnerable subgroups of children who experience multiple interacting adversities already during childhood.

*Implications of the available evidence:* Childhood adversity is a multi-layered phenomenon that cannot be fully understood through single-domain analyses. Our findings suggest that public health and social policies must address adversity more broadly, considering the interplay between individual vulnerability, family adversity, and the structural conditions of the communities in which children grow up. These insights can inform more targeted interventions; from reducing premature births and supporting families in adversity to promoting mental health and tackling broader social determinants such as poverty, education, and neighborhood conditions. By adopting a multi-layered, life course approach to childhood adversity, health and social systems can better identify and support highly vulnerable subgroups of children facing adversity, ultimately reducing intergenerational cycles of disadvantage.

## INTRODUCTION

Reducing health inequality remains a critical public health priority.^1,2^ The causal drivers of inequality are multifaceted and dynamic, involving multiple factors operating and interacting across different layers over the lifecourse.^3^ Childhood is a particularly sensitive period marked by physical, social, and cognitive development. Experiencing family-related adversity during childhood, such as poverty, disease or loss in the family, or growing up in a dysfunctional household characterized by for example parental mental health or substance abuse, has lifelong health consequences.^4–8^ While research on childhood adversity has largely focused on the family, some argue for a broader perspective that considers the child’s wider environment.^9–12^

The complexity of a child’s social environment has long been recognized,^13^ and recent frameworks increasingly emphasize that childhood adversity can emerge across multiple layers.^9–12^ This includes adversity within broader ecological contexts, such as neighborhoods.^9,14^ Living in a deprived neighborhood may restrict access to adequate health care services, healthy food environments, and safe recreational spaces. It may also induce chronic psychological stress due to a perception of threat and lack of resources, which can eventually cause physiological changes and impair health.^15^ Family adversity, for example may also interact with individual vulnerabilities, such as biological susceptibility arising from being born preterm or small-for-gestational-age.^16^ Studies show that some children face ongoing family adversity alongside poor mental and physical health throughout childhood.^4^ Using a complex systems perspective, we envision the emergence of broader patterns of adversity spanning multiple layers as the result of cascading effects, in which individual health, family social conditions, and neighborhood disadvantage act as both causes and consequences of one another throughout childhoods and across generations, ultimately contributing to lifelong disadvantage.^17^

Although death in young adulthood is rare and unexpected, it represents the most definitive public health outcome. In high-income settings, leading causes of death in young adulthood include accidents and suicide followed by, all of which are socially patterned and influenced by early-life adversity.^5^ This underscores the critical need to examine how childhood adversity across multiple layers influence later mortality risk.

Testing these cross-layer interactions requires large-scale, multilayered life course data. While several multi-level frameworks of childhood adversity exist,^9–12^ they have yet to be fully operationalized and empirically validated. In this study, we aim to investigate how multiple layers of childhood adversity relate to the risk of death in young adulthood. We hypothesize that perinatal adversity, mental and physical health adversities, and neighborhood deprivation co-occur and interact among individuals experiencing family adversity leading to higher mortality in young adulthood. To test these hypotheses, we use longitudinal, multi-layered data from a nationwide cohort of 1.2 million individuals followed from birth up to age 42.

## METHODS

### The Danish Life Course (DANLIFE) cohort

The study builds on the large register-based DANLIFE cohort^18^, which includes all children born in Denmark between 1980 and 2015, totaling 2,223,927 children. A unique personal identification number enables individual-level linkage across registries, allowing integration of adversity data across multiple layers. For this study, DANLIFE has been expanded to include information that allows identifying the neighborhood characteristics in which individuals grew up.^19^ The analysis is restricted to individuals who can be followed for entire childhoods (birth years 1980 to 2001) with complete information on adversities across all layers (n=1,235,519), see supplementary Figure S1.

Statistics Denmark and the Danish Health Authority granted access to pseudonymized data from the Danish administrative and health registers. The extended version of DANLIFE has been approved by the Danish Data Protection Agency through the records of research projects that involve personal data at the University of Copenhagen (record no 514-0641/21-3000).

### The multiple layers of childhood adversity

We conceptualize childhood adversity as occurring across two life-course dimensions (Figure 1):

1. Time: Annual data from birth to 16 years of age
2. Layers: Individual, family and neighborhood

**Figure 1.**
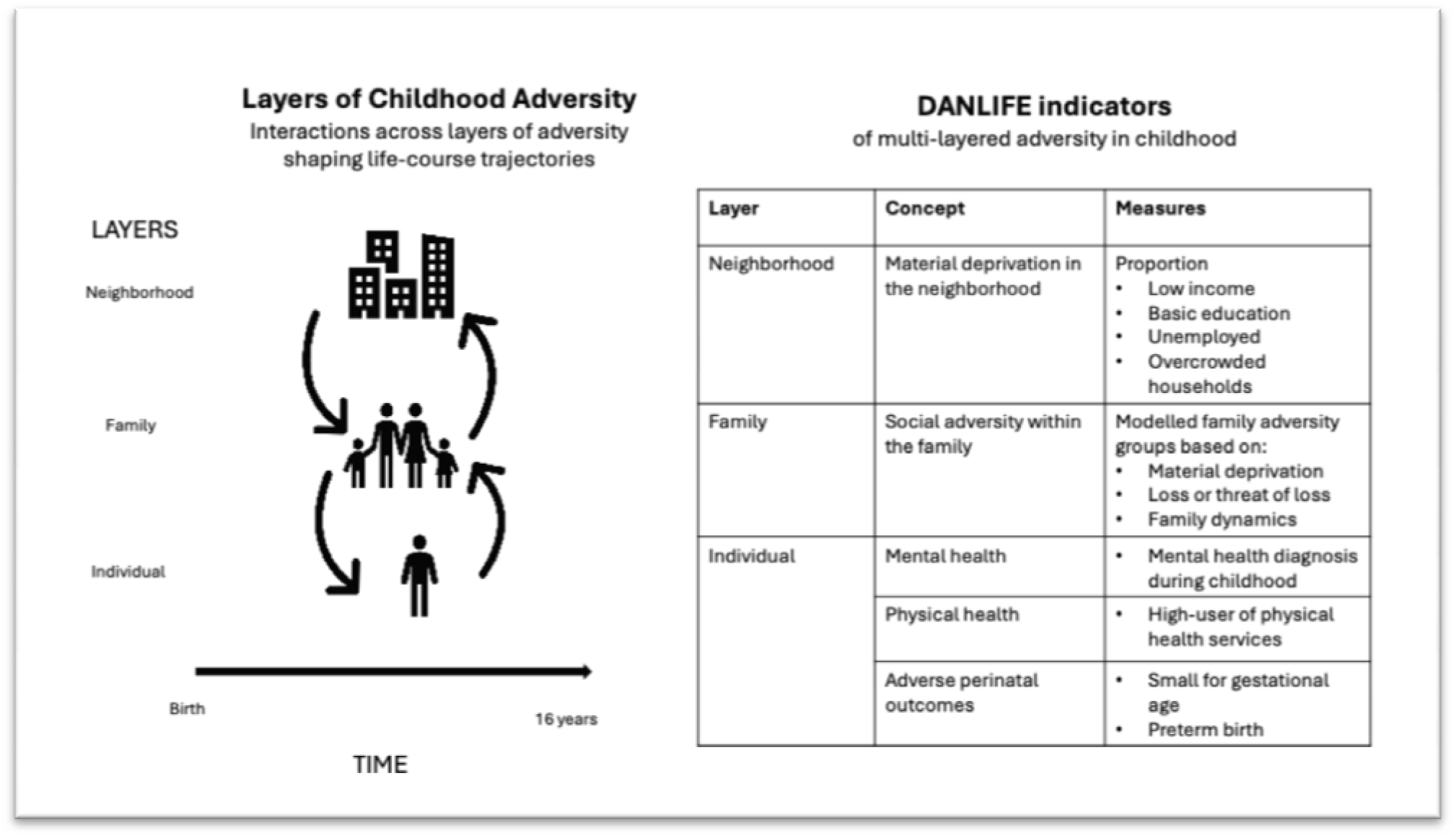
Overview of layers and measures of childhood adversity in the study.

While it is not feasible to include every aspect of childhood adversity, we selected key indicators for each layer:

i. **Individual layer.** We assessed three indicators of individual adversity in childhood:

a. *Perinatal adversity*: Defined as being born preterm (before 37 weeks of gestation) or small-for-gestational age (below the 10th percentile on age- and sex-specific growth curves^20^), based on Danish Medical Birth Register.
b. *Physical health:* Using the National Patient Registry, we calculated total emergency and inpatient hospital contacts during childhood. High physical health service use was defined as being in the top 20% for hospital encounters accumulated from age 0-15.
c. *Mental health:* Any mental health diagnosis before age 16, recorded in the Danish Psychiatric Central Research Register.
ii. **Family layer.** The family adversity layer in DANLIFE has previously been established, using register linkages between children, parents, and siblings to capture a range of social adversities in five groups.^5^ An expert panel identified three key dimensions: material deprivation (family poverty, parental long-term unemployment), loss or threat of loss (serious illness or death of a parent or sibling), and family dynamics (maternal separation, foster care placement, parental or sibling psychiatric illness, and parental alcohol or drug abuse). Using group-based multi-trajectory modeling of annual adversity counts across these dimensions (assuming equal weights across adversities within the same dimension) from ages 0-15, five family adversity trajectory groups were identified: low adversity (experiencing no or few adversities), early material deprivation (experiencing poverty or parental long-term unemployment before school age), persistent material deprivation (experiencing poverty or parental long-term unemployment during entire childhood), loss or threat of loss (experiencing serious illness or death of a parent or sibling), and high adversity (experiencing high and increasing levels of adversity across all three dimensions). Individuals were assigned to the trajectory group for which they had the highest probability of membership. Further details are available in the supplementary material (Table S1, Figure S2 and method description) and in Rod et al., 2020. ^5^
iii. **Neighborhood layer.** We built on previously generated data zones which represent small area geographic neighborhoods of approximately 2500 inhabitants across Denmark.^19^ These 1,885 zones, nested within 98 municipalities, were created using residential coordinates and a clustering algorithm (see Pedersen et al., 2022 for details).^19^ Neighborhood material deprivation was assessed using four annual indicators (proportion of individuals aged ≥25 years per zone):

a. Low income (lowest gross income quartile by year, sex and year of birth)
b. Basic education (primary schooling only, ≤7 years if born pre-1958; ≤9 years if born from 1958).
c. Unemployed
d. Overcrowding (households with >1 person per room)

These measures were calculated annually from 1981 to 2014, using weighted averages from 5-year bands. Values for 1980 were set equal to those from the 1981-1984 band and values for 2015–2017 were set equal to those from the 2010-2014 band to ensure full childhood coverage under the assumption that neighborhood conditions change slowly year to year. For each individual, we calculated average exposure to each indicator across childhood. High exposure was defined as being in the top 20% for any indicator. High neighborhood deprivation was defined as being in the top 20% for at least two indicators.

### All-cause mortality

All individuals were followed up from age 16 until death, emigration, or end of follow-up on December 31, 2022, with mortality as the outcome of interest. Individuals who emigrated during follow-up were censored at age of emigration. Maximum follow-up was 26 years, with a maximum age of 42 years at end of follow-up.

### Covariates

Covariates included sex assigned at birth (male/female), birth year, maternal age at time of birth (<20 years/20-30 years/>30 years) and parental country of origin. Parental country of origin was defined as Western if one or both parents were of European, North American, Australian, or New Zealand descent and non-Western if both parents are from elsewhere.

### Statistical analysis

First, we characterized the patterns of adversities across the different layers. For each of the five family adversity groups, we determined the percentage of individuals who also experienced high adversity at the individual and neighborhood layers. We visualized these patterns as a combined radar plot and as bar plots for each specific type of adversity.

Our primary analysis consisted of three steps:

1. We tested associations between each layer of adversity and mortality separately. To do so, we estimated hazard ratios (HR) using Cox proportional hazard models from age 16. These relative effects enable comparison with findings from previous studies. We complemented these analyses with estimates of hazard differences (HD) from Aalen’s additive hazards models to quantify the absolute magnitude of effects, which is directly relevant to public health policy.
2. We assessed additive cross-layer interactions between the family layer and both individual and neighborhood layers in relation to mortality. We used Aalen’s additive hazards models, which allowed for the detection of deviation from additivity of absolute effects to identify subgroups where most cases of excess mortality could potentially be prevented.^21^
3. We explored cumulative effects of adversities across the five family adversity groups and according to additional layers of either individual adversity only, neighborhood adversity only or both. We presented this as survival plots based on the Aalen Johansen model adjusting for emigration as competing risk and 95% CI for each added layer of adversity. We also fitted both Cox hazard models and Aalen’s additive models from age 16 to estimate HRs, HDs and 95% CIs of mortality in each of these groups. The reference group was those with low family adversity and no other measured adversities.

All models were adjusted by sex, birth year, maternal age at birth and parental country of origin, with supplementary analyses stratified by sex.

Two sensitivity analyses were performed. First, we added parental education (≤10 years, 10–12 years, >12 years) as a covariate, as it is difficult to disentangle parental education and material deprivation. Second, we restricted the sample to individuals born between 1981 and 1998 to exclude those for whom neighborhood data were extrapolated. We further limited the sample to births from 1994–2001, corresponding to Denmark’s transition to the ICD-10 system. These restricted analyses yielded similar results, supporting the robustness of our findings (results not shown).

All data manipulation, visualization, and statistical analyses were performed in R (version 4.4.1).^22^ More information about the specific package used for the analyses can be found in the supplementary methods.

## RESULTS

### Co-occurrence of adversity across layers

The patterns of multilayered adversity are summarized in Figure 2. The results are organized around the family layer. We find a clear pattern of family adversity being related to co-occuring adversities within all other layers (Panel B). The high adversity grouphas the highest proportion born small for gestational age, with childhood mental health diagnoses , and with high users of hospital services for physical conditions, whereas the persistent material deprivation group has the highest proportion living in deprived neighborhood (Panel A). Details about the distribution of covariates can be found in supplementary Table S2.

**Figure 2.**
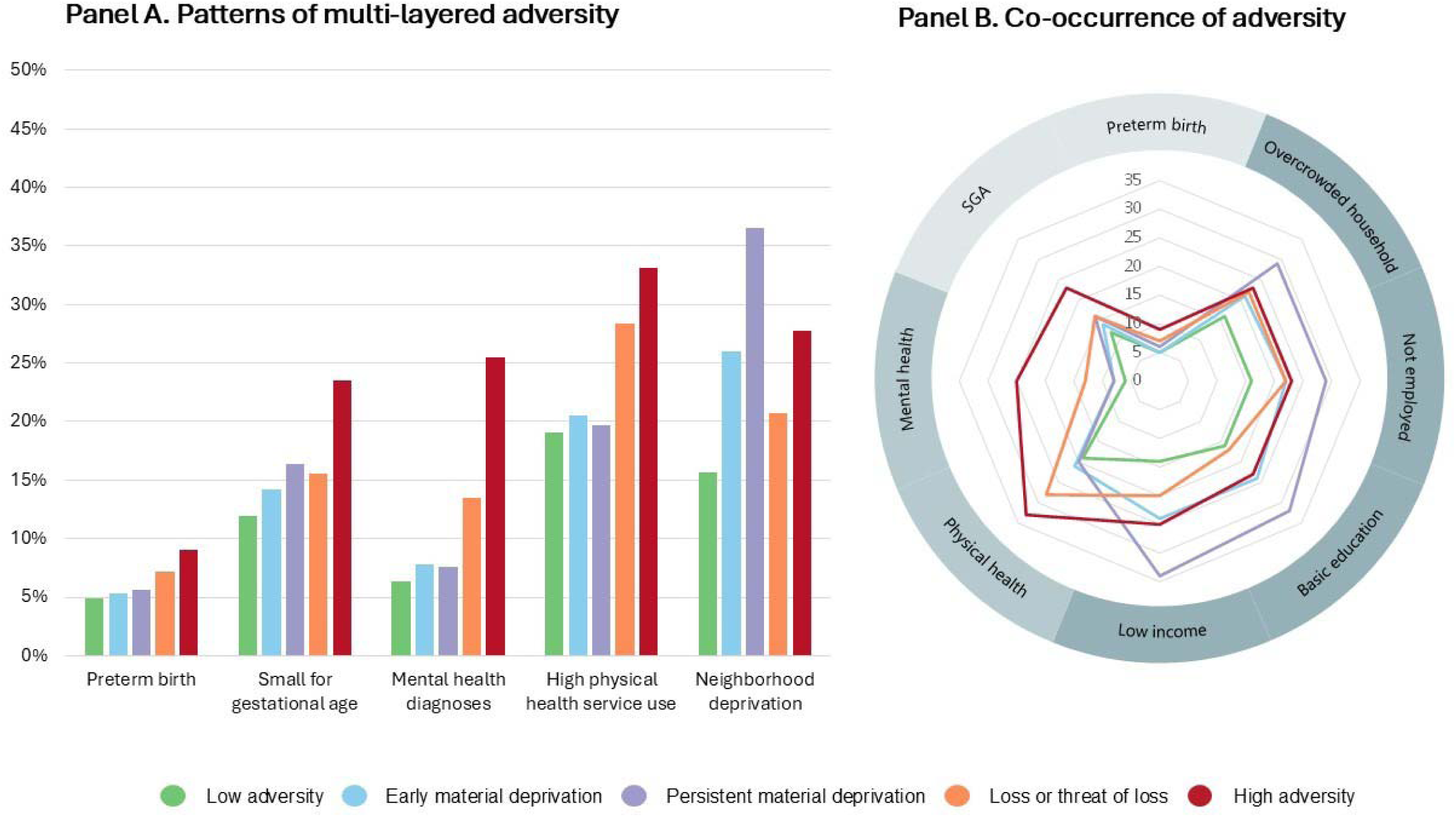
Petterns of multi-layered adversity (Panel A) and co-occurrence of childhood adversity across multiple layers of adversity according to trajectory groups of family adversity.

### Single-layer adversity and mortality

We recorded 7,320 deaths during a mean follow-up time of 14.4 years (SD = 6.4). The leading causes of death included suicide and assault (n=1321), accidents (n=1291), and cancer (n=1183). We find that each layer of adversity is associated with mortality when analyzed separately (Figure 3). For the individual layer, we show that *perinatal adversity* (hazard ratio (HR): 1.36; 95% CI: 1.28; 1.43), a *mental health diagnosis* (HR: 3.00; 95% CI: 2.82; 3.19), or being a *high user of hospital services* for physical conditions (HR: 2.36.; 95% CI: 2.24; 2.48) during childhood are associated with a higher risk of death.

**Figure 3.**
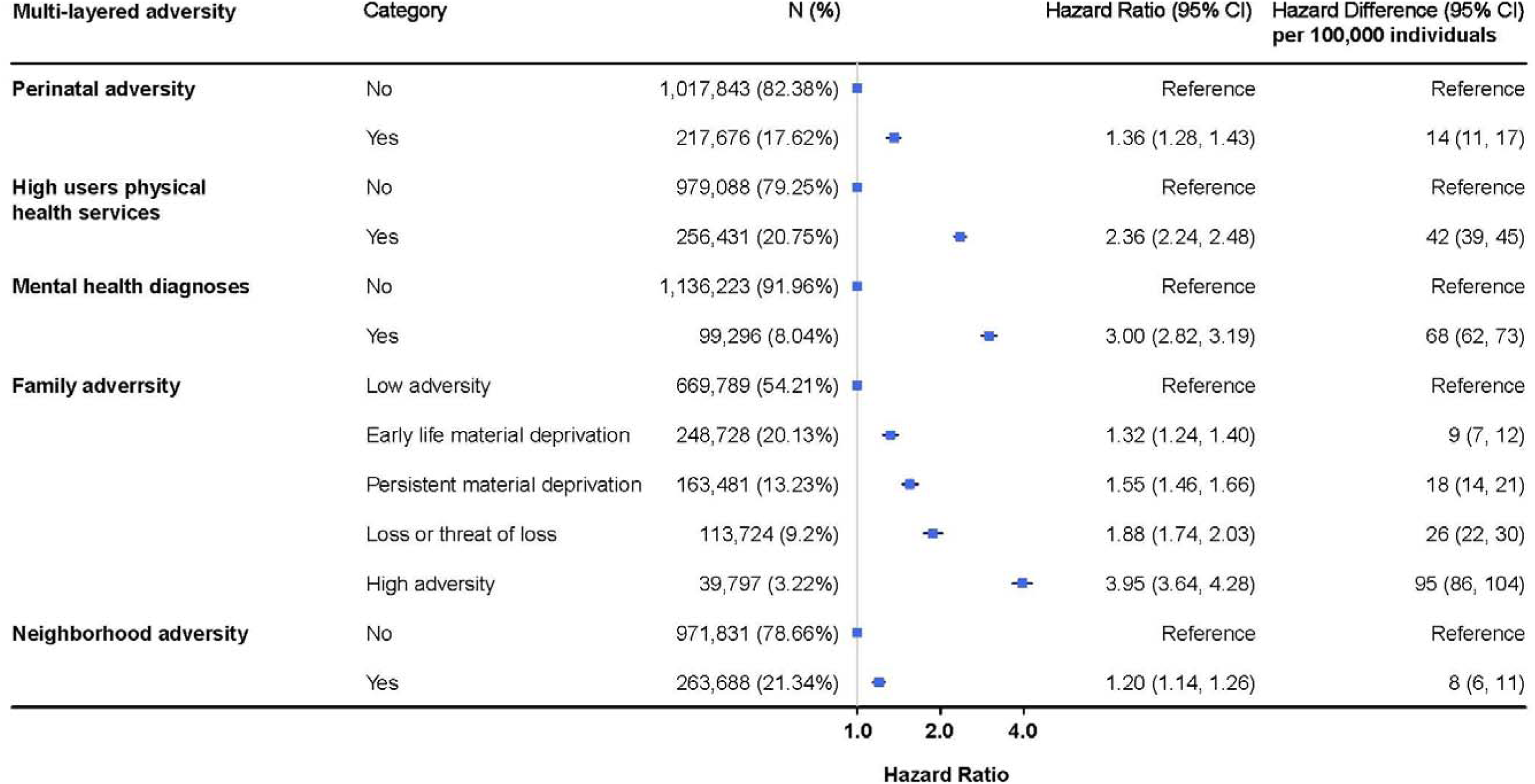
Single-layer adversity and mortality. Adjusted hazard ratios (aHR) and 95% CI for all-cause mortality in 1,235,519 young adults (16 to 42 years) according to each single layer of childhood adversity. Adjusted for sex assigned at birth, maternal age at birth, parental country of origin, and birth year.

For the family layer and in line with our previous findings,^5^ we show that trajectories of *family adversity* are associated with higher mortality.For example, high versus low adversity within the family is associated with a HR of 3.95 (95% CI: 3.64; 4.28).

For the neighborhood layer, we show that living in neighborhoods with *high material deprivation* (HR: 1.20; 95% CI: 1.14; 1.26) during childhood is associated with a higher mortality risk.

### Interactions between the family and individual layer

We find cross-layer interactions between the family and individual layer, meaning that more deaths are observed than what would have been expected by the sum of adversity at each single layer (Figure 4, supplementary Table S3). We find that those who are exposed to both early *perinatal adversity* and high family adversity experience 116 additional deaths (95% CI: 98 to 134) per 100,000 individuals compared to those with low family adversity and no perinatal adversity. Of these, 22 are due to cross-layer interaction (P=0.02). We also find that those who have both a *mental health diagnosis* and high family adversity during childhood show 181 additional deaths (95% CI: 157 to 205) per 100,000 individuals compared to those with low adversity and no mental health diagnosis. Of these, 64 are due to interaction between adversity across layers (P<.0001). In addition, we find that those who are high users of *physical health* service and experience high family adversity show 194 additional deaths (95% CI: 172 to 216) per 100,000 individuals compared to those with low adversity at both layers. Of these, an estimated 106 cases are due to cross-layer interaction (P<.0001)

**Figure 4.**
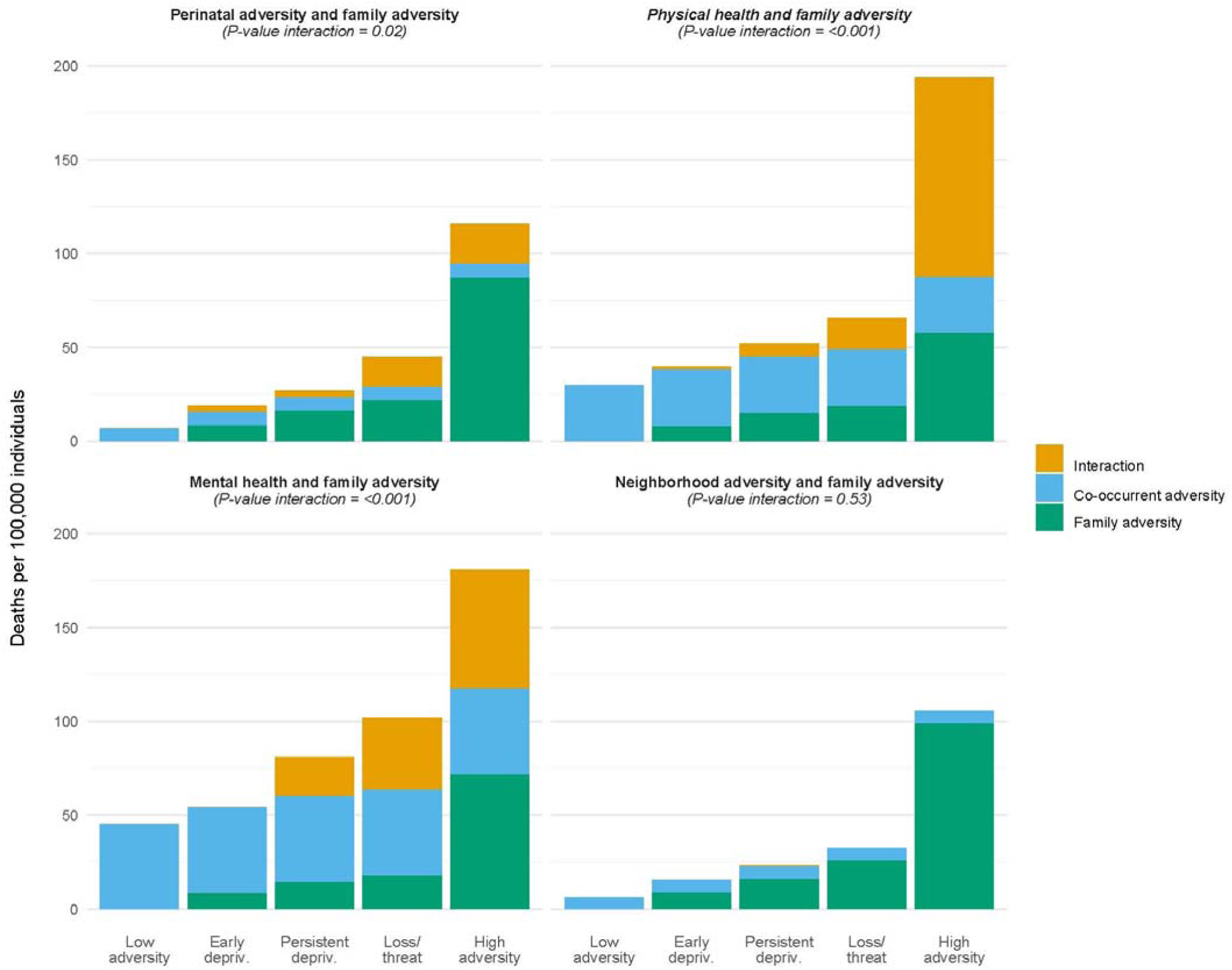
Cross-layer interactions. Adjusted hazard differences (aHD) and 95% CI for all-cause mortality in 1,235,519 young adults (16 to 42 years) according to interactions between family adversity and perinatal adversity, high user of physical health services, mental health diagnosis, and neighborhood material deprivation. Adjusted for sex assigned at birth, maternal age at birth, parental country of origin, and birth year.

### Interactions between the family and neighborhood layer

We find no interaction between the family and neighborhood layer (Figure 4, supplementary Table S3). However, individuals exposed to both high family and neighborhood adversity have a higher risk than the reference group, with 89 additional deaths (95% CI: 72–105) per 100,000 individuals due to their independent effects.

### Cumulative adversity across layers and mortality

Figure 5 shows the added mortality risk from the individual and neighborhood layer among individuals in each of the five family adversity groups. The individual layer is defined as having at least one of the three indicators (perinatal adversity, high physical health service use or mental health diagnosis). All analyses use the risk among those with low family adversity and no adversity across the other two layers as reference (367,002 individuals; 29.7% of sample). We find that adversity across multiple layers is associated with a higher mortality risk across all family adversity groups. For example, those who experience both individual adversity and neighborhood adversity despite being in the low family adversity group, still experience a higher mortality risk (HR: 1.82; 95% CI: 1.67; 1.98) than those with low adversity across layers. The highest risk is, however, found among those in the high family adversity group who also experience individual layer adversity (HR: 7.16; 95% CI: 6.40; 8.01) compared to those with low adversity. This group includes 17,810 individuals (1.4% of the population) with 434 deaths. Supplementary Table S4. shows estimates across all different combinations of adversity layers.

**Figure 5.**
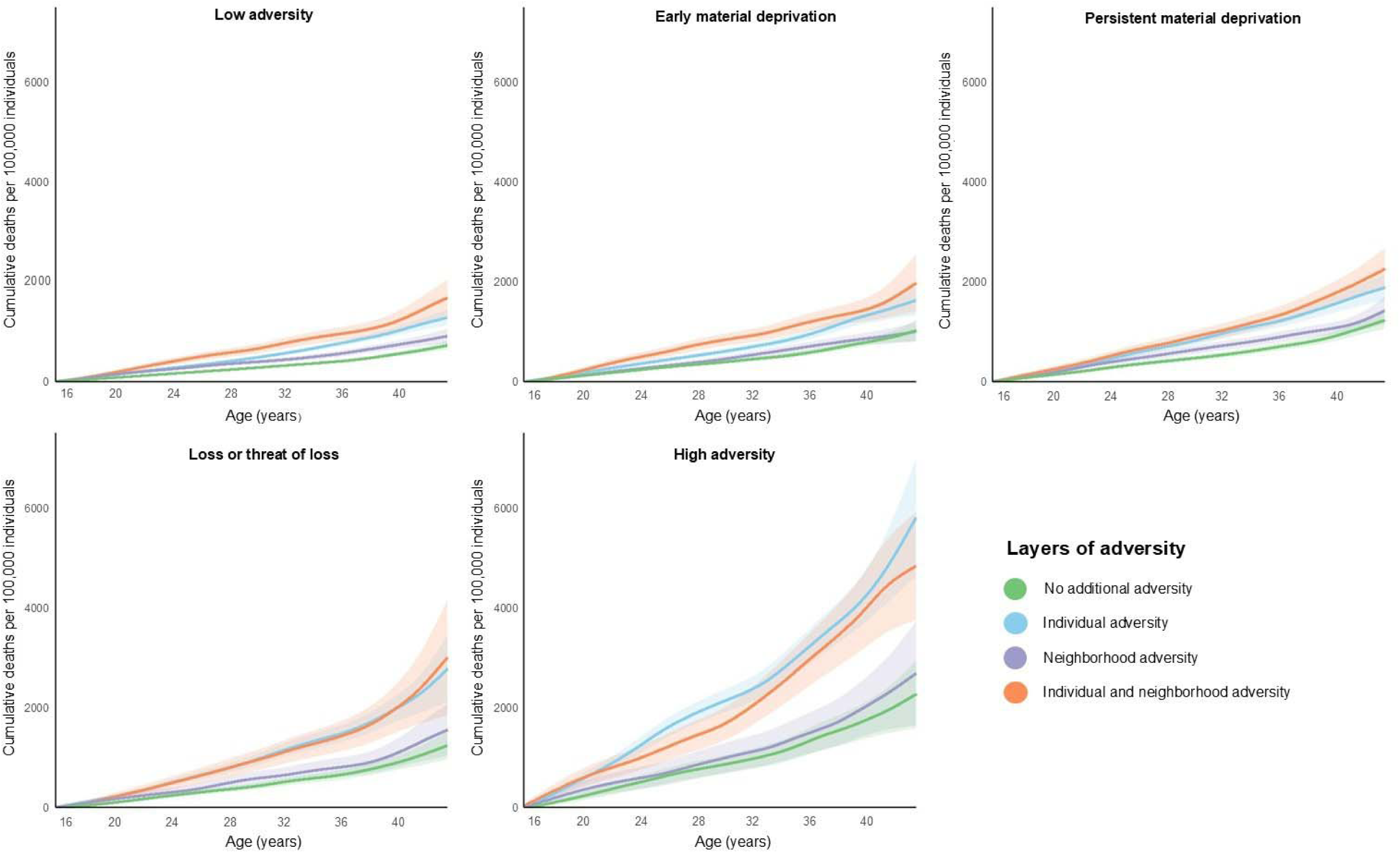
Cumulative adversity and mortality. Survival plots and 95% CI for all-cause mortality across the five family adversity groups and according to additional layers of either individual adversity only, neighborhood adversity only or both. The individual layer is defined as having one or more of the three indicators (perinatal adversity, high physical health service use or mental health diagnosis). Adjusted for sex assigned at birth, maternal age at birth, parental country of origin, and birth year.

Mortality was higher in men (5,126 deaths) than in women (2,194 deaths). Although relative effects were similar by sex, absolute risks were greatest among men (see Supplementary Tables S5–S6, Figures S3–S4). Adjusting for parental education only slightly reduced effect sizes, confirming the robustness of our results (Supplementary Table S7).

## DISCUSSION

Leveraging multi-layer life course data, we empirically demonstrated a clear co-occurrence of adversity across individual, family, and neighborhood layers. Individuals experiencing high family adversity were more likely to also experience perinatal adversity, to experience mental health problems and be high users of physical health care services, and to live in deprived neighborhoods. We also show that each of these layers of adversity is not only individually associated with mortality in young adulthood but that some of these also exhibit cross-layer interactions, where adversity at one layer accentuates the harmful effects of adversity at another layer. Such cross-layer interactions were found for the joint exposure to adversity at the individual (health and perinatal adversity) and the family layer. These interactions can create vulnerable population subgroups, starting in childhood, potentially constituting a strong driver of lifelong health inequality.

Our study highlights the significance of the child’s intrafamilial environment in shaping their ability to cope and learn, aligning with previous research that links family-related childhood adversities to health-harming behaviours, chronic conditions, and mortality in adulthood.^5–7,25^ As suggested in previous studies, we find that children who suffer from individual adversity and family adversity simultaneously during childhood are at a particularly high risk of subsequent health problems. Together, our findings provide empirical support for conceptualizing adversity as a multi-layered construct that examines the complex interplay between the child and their environment.^9–12^ Prior studies have recognized the multi-layered nature of adversity and tested indicators of childhood adversity across various layers.^23,24,26^ For example, a UK study using data from 19,000 children identified latent profiles spanning individual, family, and environmental layers and linked these profiles to developmental outcomes in adolescence.^23^ Consistent with our findings, they observed that exposure to multiple forms of early adversity is associated with a higher risk of negative outcomes in adolescence. We extend these findings to a much larger sample, enabling direct assessment of cross-scale interactions. Additionally, we utilize data spanning over four decades to track mortality into adult life.

Our findings also underscore the importance of neighborhood factors in shaping health outcomes. Although experiencing neighborhood deprivation is less associated with mortality than adversity at other layers, the neighborhood nevertheless plays a role for mortality. Most children spend the majority of their time within small geographical areas, and there is growing interest in understanding how living in certain neighborhoods can negatively impact health and contribute to health inequalities.^15^ A study utilizing register data on socioeconomic indicators at both the family and neighborhood levels found that individuals of low socioeconomic status living in deprived neighborhoods faced a higher risk of self-harm and violent crime compared with those in more affluent areas.^24^ Building on these findings, our study demonstrates how neighborhood material deprivation co-occur with adversity within families and independently affects mortality. Although neighbourhood adversity did not seem to interact with family adversity, these results still highlight the need for policies aimed at addressing neighborhood deprivation in addition to other types of adversity. Such policies could include interventions targeting housing, living conditions, poverty, and crime prevention.^27^ Future research should explore the dynamic nature of adversity, for example by examing how individual and family adversity changes due to residential mobility, offering insight into how adversity unfolds and when interventions may be most effective.

The vulnerability we document here stems from a combination of differences in exposure, susceptibility, and most likely also capacity to respond to adversity.^28^ While family adversity shows the strongest association with mortality, our findings indicate that all layers of adversity present risk, even among children who are not exposed to family adversity. Preventive strategies should extend beyond addressing only individuals at high risk and also target vulnerable subgroups experiencing disadvantages in any adversity layers. This approach requires expanding services from an individual focus to delivering integrated support for entire families who face complex needs.^29^ Policies should prioritize building contextual resources - such as social support, institutional responsiveness and access to services - to strengthen the ability of children, families and neighborhoods with adversity to adapt and recover, supporting multi-level interventions that increase health equity by expanding options, not just reducing exposures.^30^

### Strength and limitations

The multi-layered data infrastructure covering almost complete national life course data over four decades in more than 1.2 million individuals provided unique opportunities for exploring interactions between adversities across layers. Further, the study benefited from previous Danish work on small-area level neighborhoods for a detailed characterization of neighborhoods.^26^ We have focused on neighborhood material deprivation in the current study, but future studies can expand this to also include measures of e.g., social fragmentation and marginalisation.

We operationalized adversity based on selected key indicators. These indicators were carefully selected based on the social ecological model and the bio-exposome model,^9,13^ but we were limited by the availability of data in the registers. For example, information on undiagnosed health conditions, housing conditions, or neighborhood physical environment were not available through Danish registries. While the most severe cases of child abuse and neglect were likely captured through foster care records, we lacked information on unreported instances occurring within families. We were also unable to include adversity measures related to broader ecological contexts, e.g., air pollution, racism, bullying, and economic crises. While we believe that our approach allows for a comprehensive examination of childhood adversity across relevant layers, we acknowledge that we may not have captured all important dimensions of childhood adversity leading to an underestimation of effects.

We focused on all-cause mortality as the most definitive public health outcome in this young population. Utilizing the linked nationwide data largely mitigated selection bias. However, we excluded children who died before age 16, some of whose deaths were likely linked to perinatal adversity. Although the child mortality rate in Denmark is very low, this exclusion likely caused an underestimation of the total effect of childhood adversity on mortality. The most common causes of death contributing to the observed mortality pattern were suicide, accidents, and cancer. While previous studies have documented associations between childhood adversity and suicide, fewer have examined accident- and cancer-related mortality in this context. Future research should explore in greater detail the underlying mechanisms, disease trajectories, and cause-specific mortality patterns, including how factors across layers cause and reinforce each other, to better understand the overall mortality pattern identified in the present study.

Denmark’s strong social security system may limit the generalizability of these findings to other socio-political contexts. Thus, our results likely represent a ‘lower bound scenario’ within a setting with free, universal health care. These findings highlight the childhood origins of health inequality and emphasize the need for upstream structural interventions to prevent adversity and its health consequences across different countries.

### Conclusion and implications

Addressing childhood adversity requires a comprehensive, multi-layered approach. We show that multiple adversities across individual, family, and neighborhood layers intersect, trapping some children in cycles of disadvantage and widening health inequalities. Our findings highlight the need for health inequality research and interventions that address the full spectrum of adversity. Overlooking these interconnected factors carries the risk of oversimplifying complex health issues and potentially limiting intervention effectiveness. By recognizing that childhood adversity goes beyond the family context, we can integrate policies across multiple levels – including policies to reduce premature births, support disadvantaged families, promote mental health, and address broader social determinants like poverty, education, and neighborhood characteristics - to more effectively improve outcomes for children and families in vulnerable positions.

## Data Availability

It is not possible to share the individual participant data used for this study. Inquiries about secure access to the DANLIFE data under conditions stipulated by the Danish Data Protection Agency can be directed to the principal investigator of the study, Naja Hulvej Rod.

## Author contributions

NHR, SKB, LKE, AGZ and TRV conceived the idea for the study and designed the study. DK and TRV did data linkage and data cleaning. SKB, LKE, AGZ, and TRV performed the analyses for the study. NHR, SKB, LKE, DK, AGZ and TR had access to all study data. NHR wrote the first and final draft of the manuscript. All authors discussed the results and implications of the study. All authors reviewed the manuscript and have seen and approved the final manuscript.

## Data sharing statement

Individual participant data used in this study is not publicly available. Inquiries about secure access to the DANLIFE data under conditions stipulated by the Danish Data Protection Agency can be directed to principal investigator of the study Naja Hulvej Rod. An interactive map of the data zones can be found online (ncrr-au.shinyapps.io/data_zones/).

## Declaration of generative AI and AI-assisted technologies in the manuscript preparation process

During the preparation of this work the authors used perplexity.ai in order to improve readability and language. After using this service, the authors reviewed and edited the content as needed and take full responsibility for the content of the published article.

