## Supplementary materials for "The Multiple Layers of Childhood Adversity and Mortality: Population-based Study Synthesizing Life-course Data on Individual, Family and Neighborhood Adversity in 1.2 million individuals"

**OVERVIEW**

- **Supplementary Figure S1.** Flow chart.
- **Supplementary Table S1.** Dimensions and definitions of childhood adversities in the five trajectory groups of adversity.
- **Supplementary Figure S2.** Trajectories of childhood adversities across predefined dimensions of material deprivation, loss or threat of loss, and family dynamics presented as rates per person-year (PYR).
- **Supplementary Methods**. Group-based multi-trajectory model & Survival models
- **Supplementary Table S2.** Descriptive statistics of the study population by childhood adversity trajectory group membership and overall
- **Supplementary Table S3.** Additive interactions between the family layer, individual layer, and neighborhood layer (per 100,000 individuals)
- **Supplementary Table S4:** Cumulative mortality across family adversity groups and other layers of adversity (per 100,000 individuals)
- **Supplementary Table S5:** Cumulative mortality across family adversity groups and other layers of adversity - men only (per 100,000 individuals)
- **Supplementary Figure S3:** Cumulative mortality across family adversity groups and other layers of adversity - men only
- **Supplementary Table S6:** Cumulative mortality across family adversity groups and other layers of adversity – women only (per 100,000 individuals)
- **Supplementary Figure S4:** Cumulative mortality across family adversity groups and other layers of adversity – women only
- **Supplementary Table S7:** Cumulative mortality across family adversity groups and other layers of adversity – adjusted for parental education (per 100,000 individuals)
- **Supplementary Figure S4:** Cumulative mortality across family adversity groups and other layers of adversity – adjusted for parental education

**Supplementary Figure S1.** Flowchart

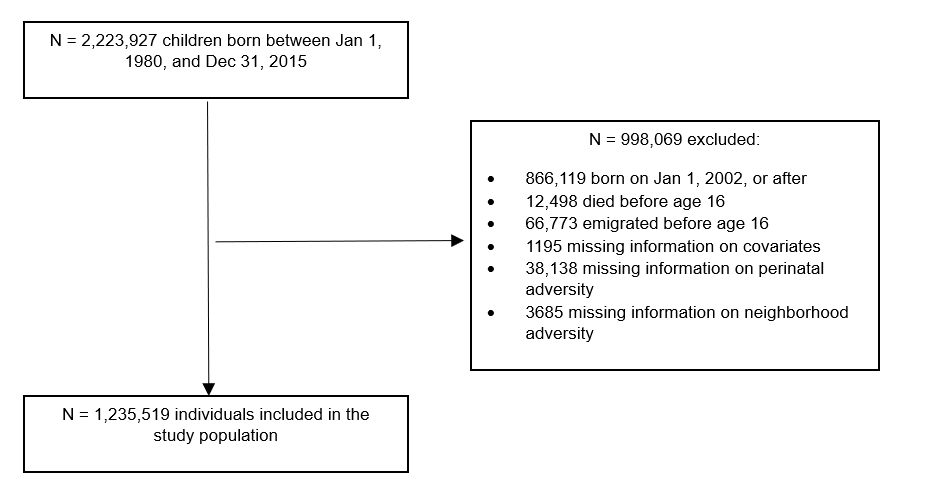

**Supplementary Table S1.** Dimensions and definitions of family adversities in the five trajectory groups of adversity identified by Rod et al. (1). Details described in Bengtsson et al. (2). The authors on Rod et al. (1) included experts in stress, child health, and child psychology and this panel of experts decided on the three predefined dimensions of childhood adversity.

|  | Adversity | Definition | Registers |
| --- | --- | --- | --- |
| Material deprivation | **Family poverty** | Family income below 50% of the median national family income in a given year | The Income Statistics Register (3) |
|  | **Parental long-term unemployment** | A parent being unemployed for at least 12 months | The Integrated Database for Labour Market Research (4) |
| Loss or threat of loss | **Death of a parent** | Death of a parent | The Danish Civil Registration System (5) |
|  | **Death of a sibling** | Death of a sibling | The Danish Civil Registration System (5) |
|  | **Parental somatic illness** | A parent being diagnosed with one of the diseases included in the Charlson comorbidity index | The Danish National Patient Register (6) |
|  | **Sibling somatic illness** | A sibling being diagnosed with one of the seven somatic illnesses most commonly related to mortality in children aged 0-18 years in Denmark: malignant neoplasm; congenital anomalies of the heart and circulatory system; congenital anomalies of the nervous system; cerebral palsy; epilepsy; cardiomyopathy; congenital disorders of lipid metabolism | The Danish National Patient Register (6) |
| Family dynamics | **Foster care** | Being placed in out-of-home care | The Register of Support for Children and Adolescents (7) |
|  | **Parental psychiatric illness** | A parent being admitted for at least 1 day to a psychiatric hospital or ward with a primary diagnosis related to psychiatric illness (excluding primary diagnoses related to alcohol and drug abuse) | The Danish Psychiatric Central Research Register (8); The Danish National Patient Register (6) |
|  | **Sibling psychiatric illness** | A sibling being admitted for at least 1 day to a psychiatric hospital or ward with a primary diagnosis related to psychiatric illness | The Danish Psychiatric Central Research Register (8); The Danish National Patient Register (6) |
|  | **Parental alcohol abuse** | A parent being diagnosed with a disease related to alcohol abuse or buying a prescribed drug used in treatment of alcohol dependence | The Danish Psychiatric Central Research Register (8); The Danish National Patient Register (6); The Danish National Prescription Registry (9) |
|  | **Parental drug abuse** | A parent being diagnosed with a disease related to drug abuse or buying a prescribed drug used in treatment of drug dependence | The Danish Psychiatric Central Research Register (8); The Danish National Patient Register (6); The Danish National Prescription Registry (9) |
|  | **Maternal separation** | The mother no longer sharing address with a partner | The Danish Civil Registration System (5) |

**Supplementary Figure S2.** Trajectories of childhood adversities across predefined dimensions of material deprivation, loss or threat of loss, and family dynamics presented as rates per person-year (PYR).

Annual rates of childhood adversity in the five trajectory groups from age 0-15 across three predefined dimensions; material deprivation (upper panels), loss or threat of loss (middle panels) and family dysfunction (lower panels). The annual rates are presented as events per person-year. The columns represent the five trajectory groups: Low Adversity (green), Early Life Material Deprivation (blue), Persistent Material Deprivation (yellow), Loss or Threat of Loss (orange) and High Adversity (red). The percentages refer to the percentage of children belonging to each trajectory group in the sample, e.g., 3% of the children belong to the high adversity group. See Supplementary Table 1 for a detailed description of the adversities included in each of the three dimensions.

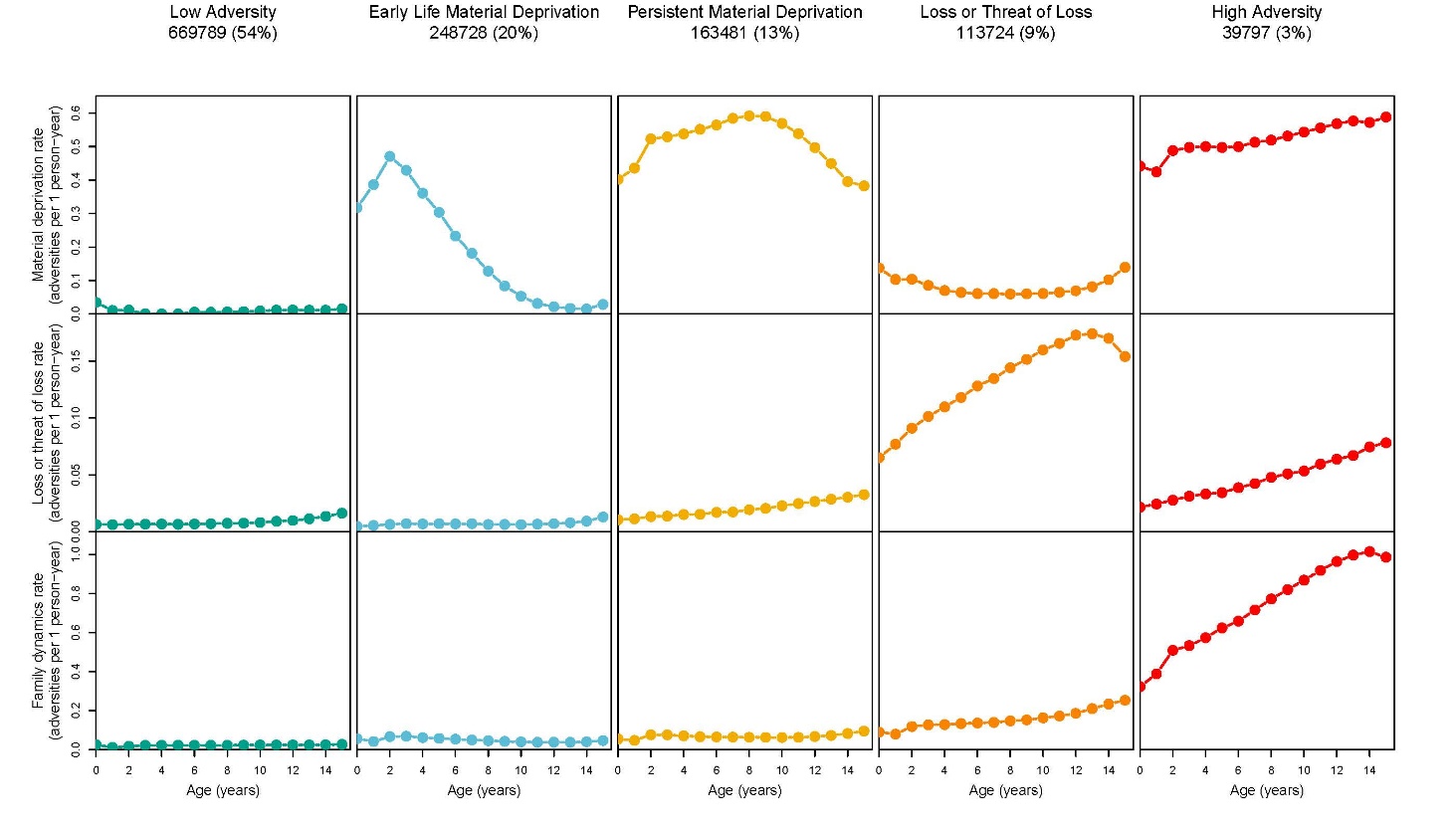

**Supplementary Methods.** Group-based multi-trajectory model & survival models

**Group-based multi-trajectory modeling**

A group-based multi-trajectory modeling was applied following the approach in Nagin, D. S., Jones, B. L., Passos, V. L., & Tremblay, R. E. (2018). Group-based multi-trajectory modeling. Statistical methods in medical research, 27(7), 2015- 2023. We aimed to determine the most common trajectory groups of family childhood adversity based on adversities in three predefined dimensions: material deprivation, loss or threat of loss, and family dynamics. For each child, we summed the number of annual family childhood adversity experiences per dimension as illustrated in the hypothetical example below depicting one imaginary child:

*Hypothetical example of one study participant*

| ***Dimension*** | ***Event type*** | ***Age 0-1*** | ***Age 1-2*** | ***(…)*** | ***Age 15-16*** |
| --- | --- | --- | --- | --- | --- |
| Material deprivation | Family poverty | 1 | 0 |  | 1 |
|  | Long-term unemployment | 0 | 2 |  | 1 |
| **Sum vector for dimension** | | **1** | **2** |  | **2** |
| Loss or threat of loss | Death of a parent | 1 | 0 |  | 0 |
|  | Death of a sibling | 0 | 1 |  | 0 |
|  | Parental somatic illness | 0 | 2 |  | 0 |
|  | Sibling somatic illness | 1 | 0 |  | 0 |
| **Sum vector for dimension 2** | | **2** | **3** |  | **0** |
| Family dynamics | Foster care | 2 | 0 |  | 0 |
|  | Parental psychiatric illness | 0 | 0 |  | 0 |
|  | Sibling psychiatric illness | 0 | 0 |  | 0 |
|  | Parental alcohol abuse | 0 | 0 |  | 0 |
|  | Parental drug abuse | 0 | 0 |  | 0 |
|  | Maternal separation | 0 | 0 |  | 0 |
| **Sum vector for dimension 3** | | **2** | **0** |  | **0** |

As outlined above, each adversity experience only counts in the year in which it occurs, but the same type of adversity can occur multiple times during childhood. As an example, see the cell in column “Age 1-2” row “Parental somatic illness”. Here, having a mother with cancer will count towards the ‘loss or threat of loss’ dimension only in the years in which the mother has a cancer diagnosis in the hospital register. Additionally, having two parents with cancer within the same year will result in a count of two in the giving year. Thus, this imaginary participant has two parents with hospital contacts for severe illnesses during his/her second year of live.

We used the package TRAJ for Stata to fit group-based multi-trajectory models using zero-inflated Poisson regressions and modeled the trajectories with a cubic function of age. The model yielded a probability for each individual of being in each trajectory group. Average posterior probabilities were 0.92, 0.82, 0.88, 0.83, and 0.94 for the low adversity, early material deprivation, persistent material deprivation, loss or threat of loss, and high adversity groups, respectively. Given these high average posterior probabilities, individuals were assigned to the cluster to which they had the highest probability of belonging (modal assignment).

Due to computational issues with large data, we employed a 2-stage approach. First, we fitted a model based on a random sample of 50,000 individuals. Second, we extrapolated the estimated probabilities of being in each trajectory group onto the full cohort. To assess the consistency of estimating the model on a sub-sample, we re-ran the procedure on 5 random samples of 50,000 individuals selected from the total study population, which returned five almost identical estimated models.

**Note:** The supplementary methods on group-based multi-trajectory modeling are replicated from the supplementary material of Rod NH, Bengtsson J, Elsenburg LK, Taylor-Robinson D, Rieckmann A. Hospitalisation patterns among children exposed to childhood adversity: a population-based cohort study of half a million children. Lancet Public Health 2021; published online Sept 29. http://dx.doi.org/10.1016/S2468-2667(21)00158-4.

**Survival models**

All analysis were conducted in R using the *survival* package (version 3.8.3, Thernau, TM, Grambsch; Thernau, 2024) for Cox proportional hazards models, *timereg* package (version 2.0.6, Scheike & Martinussen, 2006; Scheike & Zhang, 2011) for Aalen’s additive hazards models, and *prodlim* for the Aalen Johansen model (version 2025.04.08, Gerds, 2025).

**Step 1.**

We estimated associations between each singular adversity layer and all-cause mortality using both cox proportional hazard models and aalen additive hazard models. Cox proportional hazards models were fitted to estimate hazard ratios (HRs) with 95% CIs. Proportional hazards assumptions were evaluated using Schoenfeld residuals. To account for minor deviations and potential non-proportionality for birth year and sex, all models were stratified by these variables. Aalen’s additive hazards models were fitted to estimate hazard differences (HDs) with 95%. Time-dependent effects in Aalen models were inspected visually; minor deviations from linearity were observed but were judged small, so time-invariant coefficient estimates are presented. All models in this step treated exposures as fixed (time-invariant) in line with the preceding visual inspection.

**Step 2.**

Aalen’s additive hazards models were fitted to estimate hazard differences (HDs) with 95% CIs and to assess whether the combined effect of family adversity with other layers deviated from additivity. As such, we created one exposure variable combing all adversity combinations (e.g. low family adversity and no perinatal adversity, high adversity and perinatal adversity) and entered this variable as main predictor in the aalen additive hazard models. Time-dependent effects in Aalen models were inspected visually; minor deviations from linearity were observed but were judged small, so time-invariant coefficient estimates are presented. All models in this step treated exposures as fixed (time-invariant) in line with the preceding visual inspection.

**Step 3.**

We explored cumulative exposure across the five family adversity groups and additional layers (no additional adversity, individual adversity only, neighborhood adversity only, or both) with an Aalen Johansen model, a Cox Proportional Hazards model, and an Aalen Additive Hazards model with robust standard errors (100 samples). Cumulative incidence curves for all-cause mortality were obtained using the Aalen Johansen model, account for emigration as competing risk. Cox proportional hazards models were fitted to estimate hazard ratios (HRs) with 95% CIs. Proportional hazards assumptions were evaluated using Schoenfeld residuals. To account for minor deviations and potential non-proportionality for birth year and sex, we stratified by birth year and sex. Aalen’s additive hazards models were fitted to estimate hazard differences (HDs) with 95%. Time-dependent effects in Aalen models were inspected visually; minor deviations from linearity were observed but were judged small, so time-invariant coefficient estimates are presented. Both Cox and Aalen Additive hazards models were adjusted for family clustering (by mother ID) to account for siblings within our sample.

**Supplementary Table S2.** Descriptive statistics of the study population by childhood adversity trajectory group membership and overall

| Variable | Low adversity  N = 669,789 | Early life material deprivation  N = 248,728 | Persistent material deprivation  N = 163,481 | Loss or threat of loss  N = 113,724 | High adversity  N = 39,797 | Overall  N = 1,235,519 |
| --- | --- | --- | --- | --- | --- | --- |
| **Sex assigned at birth** |  |  |  |  |  |  |
| Female | 343,686 (51.3%) | 127,244 (51.2%) | 83,505 (51.1%) | 57,956 (51.0%) | 21,577 (54.2%) | 633,968 (51.3%) |
| Male | 326,103 (48.7%) | 121,484 (48.8%) | 79,976 (48.9%) | 55,768 (49.0%) | 18,220 (45.8%) | 601,551 (48.7%) |
| **Origin of parents** |  |  |  |  |  |  |
| Non-Western | 6,638 (1.0%) | 8,639 (3.5%) | 13,716 (8.4%) | 3,835 (3.4%) | 775 (1.9%) | 33,603 (2.7%) |
| Western | 663,151 (99.0%) | 240,089 (96.5%) | 149,765 (91.6%) | 109,889 (96.6%) | 39,022 (98.1%) | 1,201,916 (97.3%) |
| **Maternal age at birth** |  |  |  |  |  |  |
| <20 years | 5,516 (0.8%) | 8,663 (3.5%) | 11,002 (6.7%) | 3,830 (3.4%) | 4,165 (10.5%) | 33,176 (2.7%) |
| 20-30 years | 434,474 (64.9%) | 181,514 (73.0%) | 115,799 (70.8%) | 72,058 (63.4%) | 26,993 (67.8%) | 830,838 (67.2%) |
| >30 years | 229,799 (34.3%) | 58,551 (23.5%) | 36,680 (22.4%) | 37,836 (33.3%) | 8,639 (21.7%) | 371,505 (30.1%) |
| **Year of birth** |  |  |  |  |  |  |
| 1980-1984 | 112,910 (16.9%) | 44,288 (17.8%) | 49,985 (30.6%) | 16,945 (14.9%) | 7,564 (19.0%) | 231,692 (18.8%) |
| 1985-1989 | 129,802 (19.4%) | 63,427 (25.5%) | 43,857 (26.8%) | 20,554 (18.1%) | 9,676 (24.3%) | 267,316 (21.6%) |
| 1990-1994 | 162,373 (24.2%) | 73,543 (29.6%) | 34,255 (21.0%) | 28,568 (25.1%) | 10,622 (26.7%) | 309,361 (25.0%) |
| 1995-1999 | 186,861 (27.9%) | 49,777 (20.0%) | 25,742 (15.7%) | 33,937 (29.8%) | 8,810 (22.1%) | 305,127 (24.7%) |
| 2000-2001 | 77,843 (11.6%) | 17,693 (7.1%) | 9,642 (5.9%) | 13,720 (12.1%) | 3,125 (7.9%) | 122,023 (9.9%) |

| **Table S3.** Additive interactions between the family layer, individual layer, and neighborhood layer (per 100,000 individuals) | | | | | | |  |
| --- | --- | --- | --- | --- | --- | --- | --- |
|  |  | Low adversity | Early material deprivation | Persistent material deprivation | Loss or threat of loss | High adversity | P-value for interaction |
| Perinatal adversity | No | Reference | 8 (6 – 11) | 17 (13 – 20) | 22 (18 – 26) | 88 (77 – 98) |  |
|  | Yes | 7 (4 – 10) | 19 (14 – 24) | 27 (20 – 33) | 45 (36 – 54) | 116 (98 – 134) |  |
|  | Due to Interaction | 0 | 4 | 3 | 16 | 22 | 0.02 |
| High users of physical health services | No | Reference | 8 (6-11) | 15 (12-18) | 19 (15-23) | 58 (49-67) |  |
|  | Yes | 30 (26 – 34) | 40 (33-46) | 52 (44-60) | 66 (56-75) | 194 (172-216) |  |
|  | Due to Interaction | 0 | 2 | 7 | 17 | 106 | < 0.0001 |
| Mental health diagnoses | No | Reference | 9 (6 – 11) | 15 (12 – 18) | 18 (15 – 22) | 72 (63 – 81) |  |
|  | Yes | 45 (38 – 53) | 51 (40 – 61) | 81 (66 – 96) | 102 (86 – 118) | 181 (157 – 205) |  |
|  | Due to Interaction | 0 | -4 | 21 | 38 | 64 | < 0.0001 |
| Neighborhood | No | Reference | 9 (6 – 12) | 16 (13 – 20) | 26 (22 – 30) | 99 (88 – 110) |  |
|  | Yes | 6 (3 – 10) | 14 (10 – 19) | 23 (18 – 28) | 30 (22 – 38) | 89 (72 – 105) |  |
|  | Due to Interaction | 0 | -1 | 1 | -2 | -17 | 0.53 |

| **Table S4.** Cumulative mortality across family adversity groups and other layers of adversity (N = 1,235,519) | | | | | |
| --- | --- | --- | --- | --- | --- |
| **Family adversity** | **Other layers of adversity** | **Deaths** | **Deaths per 100 000 individuals per year** | **Hazard ratio (95% CI)** | **Hazard difference per 100 000 individuals per year (95% CI)** |
| **Low adversity** | No additional adversity | 1160 | 2207 | Reference | Reference |
|  | Individual adversity | 969 | 39.00 | 1.82 (1.67–1.98) | 18 (15–21) |
|  | Neighborhood adversity | 369 | 32.10 | 1.28 (1.14–1.44) | 5 (2–9) |
|  | Individual and neighborhood adversity | 247 | 52.70 | 2.25 (1.96–2.58) | 29 (22–35) |
| **Early material deprivation** | No additional adversity | 553 | 32.40 | 1.38 (1.25–1.53) | 8 (5–11) |
|  | Individual adversity | 495 | 49.90 | 2.2 (1.98–2.45) | 27 (22–31) |
|  | Neighborhood adversity | 249 | 37.20 | 1.48 (1.29–1.7) | 10 (5–15) |
|  | Individual and neighborhood adversity | 226 | 63.90 | 2.69 (2.33–3.11) | 39 (30–47) |
| **Persistent material deprivation** | No additional adversity | 417 | 40.00 | 1.53 (1.37–1.72) | 11 (7–15) |
|  | Individual adversity | 423 | 66.20 | 2.6 (2.32–2.91) | 38 (32–45) |
|  | Neighborhood adversity | 318 | 49.70 | 1.85 (1.63–2.09) | 20 (14–25) |
|  | Individual and neighborhood adversity | 272 | 74.10 | 2.82 (2.47–3.23) | 45 (36–54) |
| **Loss or threat of loss** | No additional adversity | 236 | 36.00 | 1.6 (1.39–1.84) | 14 (9–18) |
|  | Individual adversity | 409 | 75.40 | 3.51 (3.14–3.94) | 55 (47–62) |
|  | Neighborhood adversity | 85 | 46.30 | 1.94 (1.56–2.43) | 21 (12–31) |
|  | Individual and neighborhood adversity | 118 | 76.10 | 3.38 (2.79–4.08) | 53 (40–67) |
| **High adversity** | No additional adversity | 123 | 72.10 | 2.89 (2.4–3.49) | 45 (32–58) |
|  | Individual adversity | 434 | 171.50 | 7.16 (6.4–8.01) | 146 (130–162) |
|  | Neighborhood adversity | 63 | 83.30 | 3.2 (2.49–4.13) | 54 (34–75) |
|  | Individual and neighborhood adversity | 154 | 152.50 | 6.04 (5.1–7.16) | 125 (101–149) |

| **Table S5.** Cumulative mortality across family adversity groups and other layers of adversity – men (N = 633,968) | | | | | |
| --- | --- | --- | --- | --- | --- |
| **Family adversity** | **Other layers of adversity** | **Deaths** | **Deaths per 100 000 individuals per year** | **Hazard ratio (95% CI)** | **Hazard difference per 100 000 individuals per year (95% CI)** |
| **Low adversity** | No additional adversity | 790 | 30.80 | Reference | Reference |
|  | Individual adversity | 671 | 49.40 | 1.76 (1.59–1.95) | 24 (19–28) |
|  | Neighborhood adversity | 269 | 45.90 | 1.35 (1.18–1.56) | 10 (4–16) |
|  | Individual and neighborhood adversity | 169 | 67.00 | 2.17 (1.83–2.56) | 36 (26–46) |
| **Early material deprivation** | No additional adversity | 368 | 43.20 | 1.34 (1.19–1.52) | 10 (5–15) |
|  | Individual adversity | 349 | 64.80 | 2.16 (1.91–2.46) | 35 (28–42) |
|  | Neighborhood adversity | 186 | 55.50 | 1.62 (1.38–1.9) | 19 (10–27) |
|  | Individual and neighborhood adversity | 166 | 86.60 | 2.75 (2.32–3.26) | 54 (41–68) |
| **Persistent material deprivation** | No additional adversity | 293 | 56.30 | 1.58 (1.38–1.81) | 17 (10–24) |
|  | Individual adversity | 295 | 85.70 | 2.54 (2.22–2.91) | 50 (40–60) |
|  | Neighborhood adversity | 230 | 72.40 | 1.96 (1.68–2.27) | 31 (21–41) |
|  | Individual and neighborhood adversity | 192 | 96.10 | 2.76 (2.35–3.25) | 58 (44–72) |
| **Loss or threat of loss** | No additional adversity | 172 | 53.00 | 1.73 (1.46–2.04) | 22 (14–30) |
|  | Individual adversity | 280 | 96.60 | 3.4 (2.96–3.9) | 70 (58–81) |
|  | Neighborhood adversity | 65 | 72.20 | 2.21 (1.71–2.86) | 38 (20–56) |
|  | Individual and neighborhood adversity | 69 | 83.20 | 2.77 (2.17–3.55) | 53 (34–73) |
| **High adversity** | No additional adversity | 92 | 104.60 | 3.09 (2.49–3.85) | 68 (46–90) |
|  | Individual adversity | 312 | 217.50 | 6.97 (6.1–7.96) | 185 (161–209) |
|  | Neighborhood adversity | 45 | 116.90 | 3.29 (2.44–4.45) | 77 (43–111) |
|  | Individual and neighborhood adversity | 113 | 200.50 | 6.08 (4.98–7.42) | 165 (128–202) |

**Supplementary Figure S2:** Cumulative mortality across family adversity groups and other layers of adversity - men only

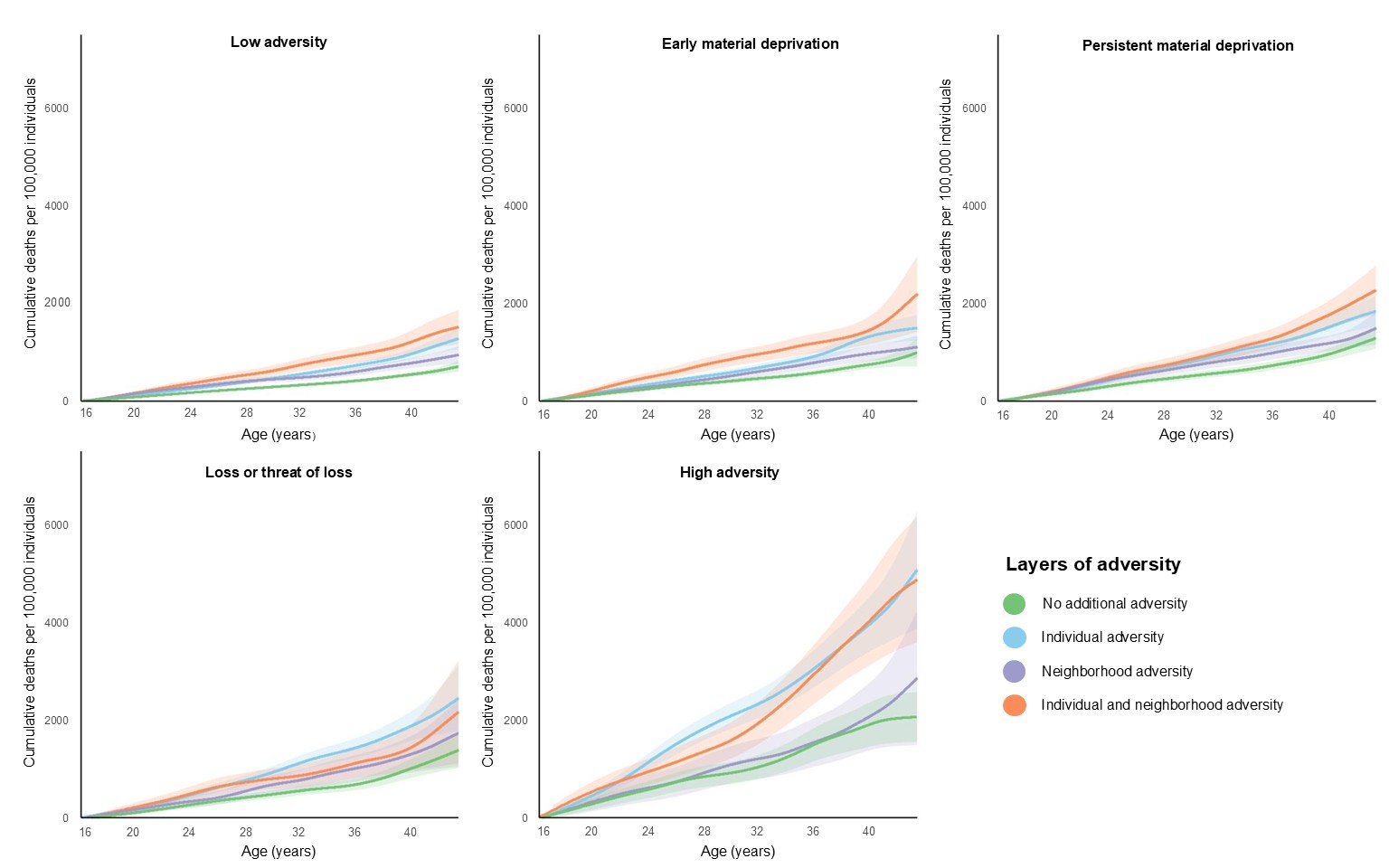

| **Table S6.** Cumulative mortality across family adversity groups and other layers of adversity – women (N = 601,551) | | | | | |
| --- | --- | --- | --- | --- | --- |
| **Family adversity** | **Other layers of adversity** | **Deaths** | **Deaths per 100 000 individuals per year** | **Hazard ratio (95% CI)** | **Hazard difference per 100 000 individuals per year (95% CI)** |
| **Low adversity** | No additional adversity | 370 | 14.50 | Reference | Reference |
|  | Individual adversity | 298 | 26.50 | 1.96 (1.68–2.28) | 14 (10–17) |
|  | Neighborhood adversity | 100 | 17.80 | 1.12 (0.9–1.39) | 1 (−3–5) |
|  | Individual and neighborhood adversity | 78 | 36.10 | 2.44 (1.91–3.11) | 21 (13–29) |
| **Early material deprivation** | No additional adversity | 185 | 21.60 | 1.45 (1.22–1.73) | 6 (3–10) |
|  | Individual adversity | 146 | 32.20 | 2.29 (1.89–2.78) | 18 (13–24) |
|  | Neighborhood adversity | 63 | 18.80 | 1.19 (0.91–1.56) | 2 (−3–7) |
|  | Individual and neighborhood adversity | 60 | 37.00 | 2.51 (1.91–3.3) | 22 (12–31) |
| **Persistent material deprivation** | No additional adversity | 124 | 23.80 | 1.44 (1.17–1.76) | 6 (1–10) |
|  | Individual adversity | 128 | 43.50 | 2.74 (2.24–3.36) | 27 (19–34) |
|  | Neighborhood adversity | 88 | 27.30 | 1.62 (1.28–2.05) | 9 (3–15) |
|  | Individual and neighborhood adversity | 80 | 47.90 | 2.95 (2.31–3.77) | 30 (20–41) |
| **Loss or threat of loss** | No additional adversity | 64 | 19.30 | 1.34 (1.02–1.75) | 5 (0–10) |
|  | Individual adversity | 129 | 51.10 | 3.79 (3.1–4.64) | 38 (29–47) |
|  | Neighborhood adversity | 20 | 21.40 | 1.39 (0.89–2.19) | 5 (−4–15) |
|  | Individual and neighborhood adversity | 49 | 67.90 | 4.85 (3.6–6.53) | 54 (35–73) |
| **High adversity** | No additional adversity | 31 | 37.50 | 2.41 (1.67–3.48) | 21 (8–34) |
|  | Individual adversity | 122 | 111.30 | 7.64 (6.2–9.41) | 96 (77–116) |
|  | Neighborhood adversity | 18 | 48.50 | 2.99 (1.86–4.8) | 31 (8–53) |
|  | Individual and neighborhood adversity | 41 | 91.80 | 5.93 (4.29–8.2) | 75 (47–103) |

**Supplementary Figure S3:** Cumulative mortality across family adversity groups and other layers of adversity - women only

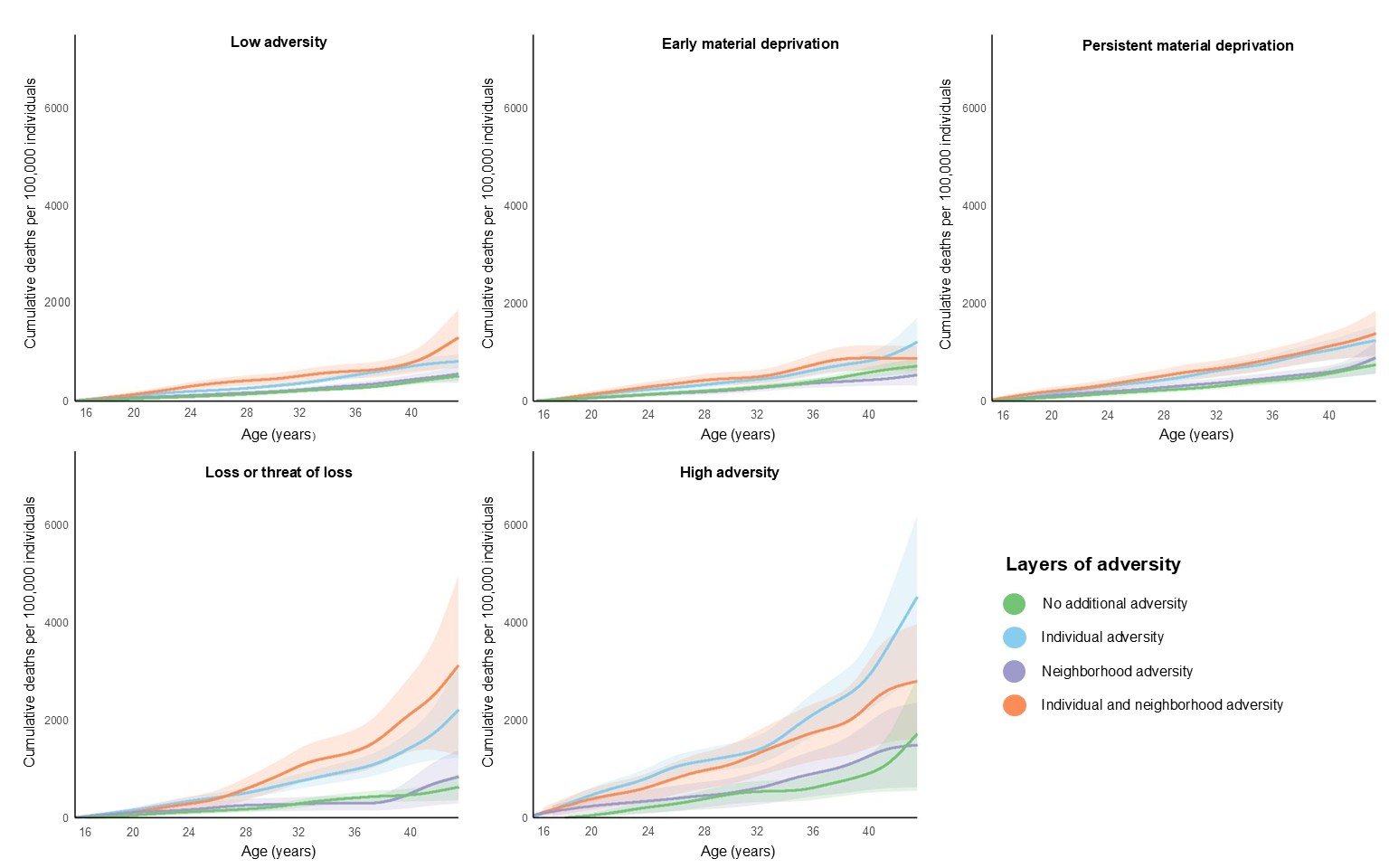

| **Table S7.** Cumulative mortality across family adversity groups and other layers of adversity – adjusted for parental educational background (N = 1,232,468) | | | | | |
| --- | --- | --- | --- | --- | --- |
| **Family adversity** | **Other layers of adversity** | **Deaths^a^** | **Deaths per 100 000 individuals per year** | **Hazard ratio (95% CI)** | **Hazard difference per 100 000 individuals per year (95% CI)** |
| **Low adversity** | No additional adversity | - | 22,70 | Reference | Reference |
|  | Individual adversity | - | 39,10 | 1.8 (1.65–1.96) | 18 (15–21) |
|  | Neighborhood adversity | - | 32,00 | 1.24 (1.1–1.4) | 4 (0–8) |
|  | Individual and neighborhood adversity | - | 52,40 | 2.14 (1.87–2.46) | 27 (20–33) |
| **Early material deprivation** | No additional adversity | - | 32,30 | 1.32 (1.19–1.46) | 6 (3–9) |
|  | Individual adversity | - | 49,80 | 2.08 (1.87–2.31) | 24 (20–29) |
|  | Neighborhood adversity | - | 37,30 | 1.4 (1.22–1.6) | 8 (3–13) |
|  | Individual and neighborhood adversity | - | 63,30 | 2.46 (2.12–2.84) | 35 (27–43) |
| **Persistent material deprivation** | No additional adversity | - | 40,10 | 2.64 (2.19–3.19) | 9 (5–13) |
|  | Individual adversity | - | 66,10 | 1.44 (1.29–1.62) | 35 (29–42) |
|  | Neighborhood adversity | - | 49,90 | 2.4 (2.14–2.69) | 16 (11–22) |
|  | Individual and neighborhood adversity | - | 74,60 | 1.7 (1.5–1.93) | 41 (32–51) |
| **Loss or threat of loss** | No additional adversity | - | 36,00 | 2.58 (2.25–2.95) | 12 (8–17) |
|  | Individual adversity | - | 75,60 | 1.55 (1.35–1.79) | 53 (45–60) |
|  | Neighborhood adversity | - | 46,00 | 3.34 (2.98–3.75) | 19 (9–29) |
|  | Individual and neighborhood adversity | - | 75,30 | 1.81 (1.45–2.27) | 49 (35–63) |
| **High adversity** | No additional adversity | - | 72,60 | 3.06 (2.52–3.71) | 42 (29–55) |
|  | Individual adversity | - | 171,30 | 6.38 (5.67–7.18) | 141 (125–157) |
|  | Neighborhood adversity | - | 82,70 | 2.83 (2.19–3.65) | 49 (28–69) |
|  | Individual and neighborhood adversity | - | 154,30 | 5.38 (4.53–6.39) | 121 (97–145) |
| **^a^** Not provided due to issues relating to identification/data privacy | | | | | |

**Supplementary Figure S5:** Cumulative mortality across family adversity groups and other layers of adversity – adjusted for parental education

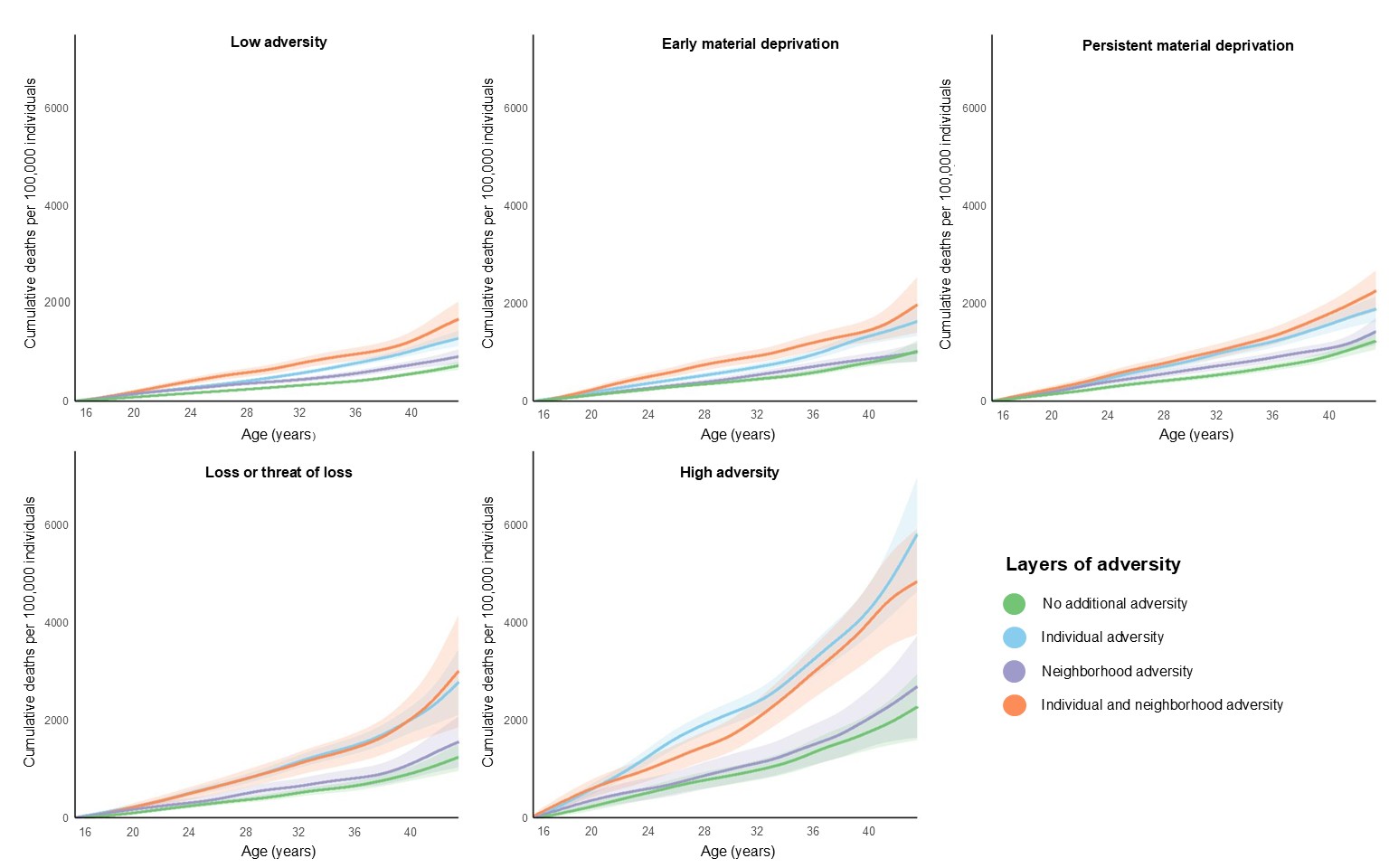
